# Knowledge, self-efficacy, and fears following a multi-station first aid workshop for health-adjacent trainees in Karachi, Pakistan: a single-arm pre–post evaluation with exploratory three-month retention

**DOI:** 10.64898/2026.06.24.26356482

**Authors:** Muhammad Rohaan Ali Jarral, Ayesha Abbasi, Shaheer Hassan Shahzad, Iqra Navroz, Yasir Shafiq, Mahwish Fatima, Aiman Shakir, Sohaib Ilyas, Amber Palla

## Abstract

Low- and middle-income countries bear over 90% of global injury-related mortality yet largely lack formal prehospital emergency systems, leaving laypersons as the primary point of care. In Pakistan, nearly 90% of out-of-hospital cardiac arrests are witnessed, but bystander resuscitation rates remain critically low, attributable to knowledge deficits, absent Good Samaritan protections, and unaddressed psychological barriers. Rigorous multi-domain evaluations of first aid training that simultaneously examine knowledge acquisition, self-efficacy, attitudinal barriers, and medium-term retention remain limited in South Asian contexts.

A single-centre, single-arm, pre-post quasi-experimental study evaluated a structured multi-station first aid workshop at a tertiary academic institution in Karachi. A convenience sample of 132 health-adjacent trainees completed six competency-based stations covering cardiopulmonary resuscitation, haemorrhage control, fracture management, foreign body airway obstruction, and burns, delivered through didactic teaching, video demonstration, mannequin-based simulation, and facilitated scenario exercises. Paired data were available for 68 participants on knowledge, 100 on self-efficacy, 65 on perceived barriers, and 15 at three-month follow-up.

Knowledge scores increased from a mean of 5.9 to 9.1 out of 13 (Cohen’s d = 1.06), a large effect. Statistically significant item-level improvements were observed in 9 of 13 items, with the greatest gains in chest-compression rate, first response to witnessed cardiac arrest, and choking management. Self-efficacy improved significantly across all seven assessed domains, with mean gains of approximately 38 to 55 points on a 100-point scale. Perceived barriers, including fear of legal liability, patient harm, and performance panic, remained unchanged. Exploratory three-month knowledge retention averaged approximately 96% among returners, although this is likely a modest upper bound, as returners scored marginally higher post-training and the comparison was underpowered.

The intervention produced substantial short-term gains in knowledge and self-efficacy. The persistence of psychological and legal barriers despite these gains suggests that cognitive instruction alone is insufficient to translate preparedness into bystander action, underscoring the need for integrated legal education and barrier-focused psychological components in future curricula.

## Introduction

The majority of the global injury burden falls on low- and middle-income countries (LMICs), where over 90% of the 4.4 million annual injury-related deaths occur [1]. In Punjab, Pakistan alone, medical emergencies comprised 66.7% of incidents managed by Rescue 1122 in 2024, totaling over 1.5 million cases [2]. In many LMICs, community members and laypersons routinely serve as de facto first responders due to the limited availability of formal emergency medical services. Replicating high-income country prehospital systems is often financially and logistically unfeasible in these settings, making context-appropriate, community-driven approaches essential [3].

First aid training represents one of the most efficient strategies to reduce the burden of injury and disease. Emergency care, including first aid, has the potential to prevent up to half of deaths and one-third of disabilities in LMICs [4]. Defined as the immediate assistance provided to a sick or injured person until professional medical help arrives, first aid encompasses actions aimed at preserving life, alleviating suffering, preventing further injury or illness, and promoting recovery [5]. Structured first aid training has consistently demonstrated effectiveness in improving knowledge, practical skills, and confidence among laypersons, students, and healthcare providers alike [6–8]. The mechanism is well-established: early recognition of cardiac arrest, prompt activation of emergency services, immediate cardiopulmonary resuscitation (CPR), and effective external bleeding control are each associated with two- to three-fold improvements in survival outcomes [9,10].

Despite this evidence, the translation of training into action remains uneven. Although nearly 90% of out-of-hospital cardiac arrests are witnessed in Pakistan, only 2.3% of victims receive bystander CPR, and overall survival is among the lowest reported globally [11]. This gap reflects multiple deficits: insufficient public exposure to structured training, low baseline knowledge, weak emergency medical services coordination, the absence of Good Samaritan legislation that would protect responders from legal exposure, and persistent psychological barriers including fear of harming the victim and reluctance to assume responsibility [11,12]. Within Pakistan, the Pakistan Life Savers Programme (PLSP), launched in 2020, aims to equip ten million citizens with hands-only CPR and bleeding control skills and has trained more than 127,000 individuals to date [11,13]. However, available evidence remains constrained: studies are heavily weighted toward CPR-only interventions, few examine self-efficacy and emotional barriers alongside knowledge, and longitudinal retention beyond the immediate post-training window is rarely reported [14]. Evaluations of comprehensive, multi-domain first aid training programs in health-adjacent populations remain conspicuously sparse in South Asian literature.

In this study, we conducted a single-centre, single-arm, pre–post quasi-experimental evaluation of a structured, multi-station first aid workshop hosted at a tertiary academic centre in Karachi and delivered to a convenience sample of health-adjacent trainees recruited from health-related programs across the city. The primary outcomes were changes in participants’ first aid knowledge and self-efficacy from immediately before to immediately after training. Secondary outcomes were changes in participants’ concerns and fears regarding the provision of first aid, and an exploratory assessment of knowledge retention at three months in a self-selected subset of participants.

## Methods

### Study Design and Setting

This quasi-experimental observational study was conducted in a series of workshops from August 2025 to January 2026 in Karachi, Pakistan. A single-arm pre-post design was employed, as withholding first aid training from a control group was not ethically justifiable or logistically feasible in this resource-limited setting where community members serve as de facto first responders. This design is standard in first aid training evaluation and aligns with the dominant methodology in the field [14].

### Study Population

A total of 132 participants received the training. Given the pragmatic, field-based nature of this evaluation, we conducted complete-case analysis for each outcome independently rather than restricting to participants with data across all instruments. This approach preserves statistical power and reflects the reality of workshop-based data collection in LMIC settings where paper-based forms are vulnerable to loss and incomplete submission. We did not assume data were missing completely at random; the sensitivity analyses in Supplementary Tables S1–S2 characterise how participants with and without complete data differ, and the affected estimates are interpreted as bounded, upper-bound values accordingly.

Sixty-eight participants had complete pre- and post-test knowledge data and were included in knowledge analysis. One hundred participitants provided complete self-efficacy data. Self-efficacy was captured on a separate, single-page instrument that was returned more consistently than the multi-page knowledge questionnaire, accounting for the higher completion for that outcome (n=100 vs n=68). Sixty-five participants completed both pre- and post-training concerns and fears assessments. Fifteen participants completed the 3-month follow-up assessment. Participants with incomplete data for a given outcome were excluded from that specific analysis but included in others where data were complete (Figure.1). A sensitivity analysis comparing baseline characteristics of participants who completed versus did not complete each assessment is presented in Supplementary Table S1.

**Figure 1.**
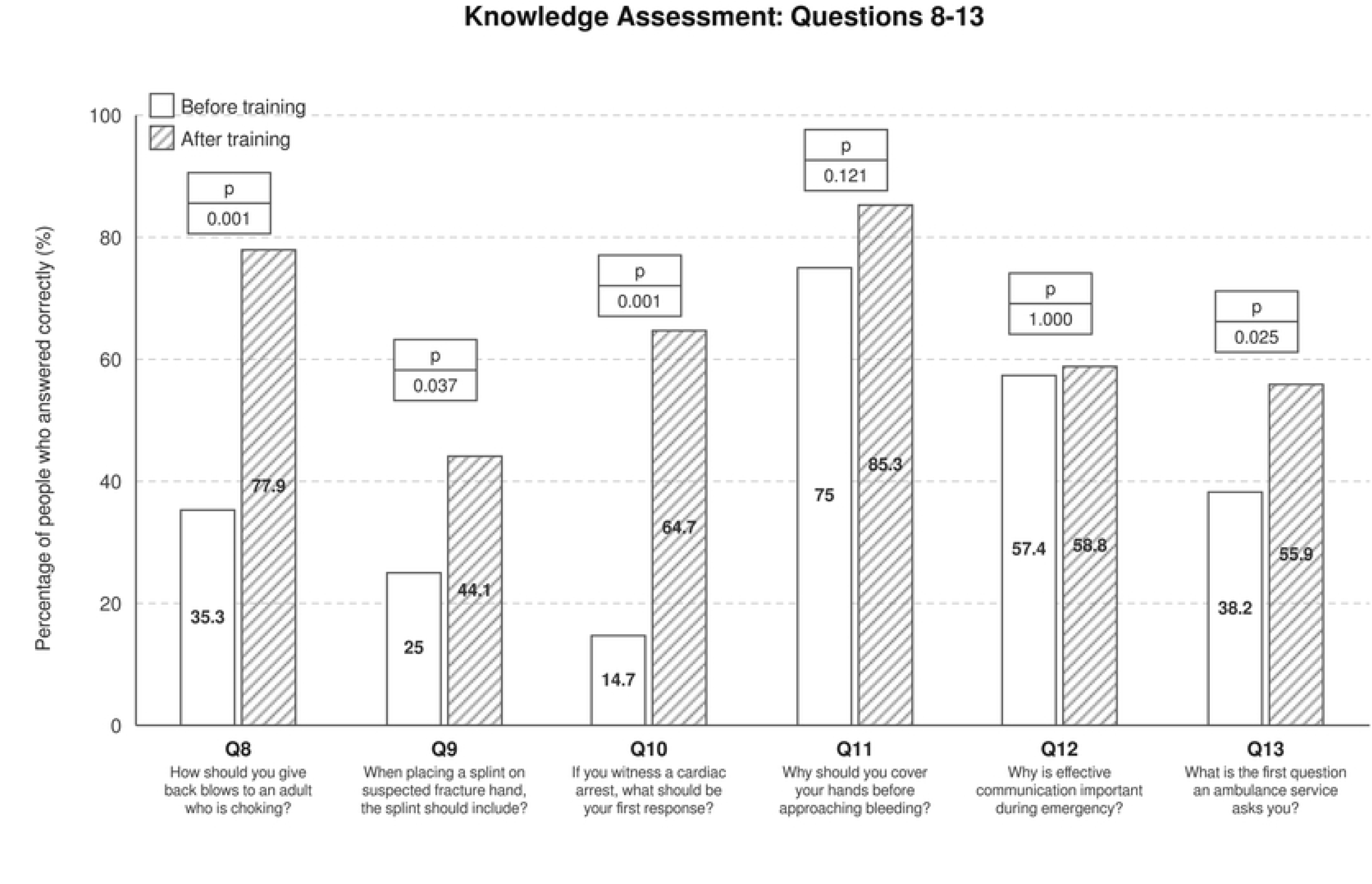
Participant flow diagram showing parallel analyses across study measures. From 132 trained participants, four independent complete-case analyses were conducted: knowledge assessment (n = 68), self-efficacy assessment (n = 100), concerns/fears assessment (n = 65), and 3-month follow-up (n = 15). Exclusion reasons are shown for each stream. These four analytic subsets derive from the single trained cohort (N = 132) and are not mutually exclusive; each outcome was analysed in all participants with complete paired data for that measure.

### Intervention and Training

Participants’ written informed consent was obtained followed by a pre-test questionnaire before being divided into small groups. Each group rotated through five skill-based training stations covering cardiopulmonary resuscitation (CPR), bleeding control, fracture management, choking and choking. Each station was facilitated by a master trainer or certified first aid responder. Facilitators provided theoretical instruction supported by video demonstrations and guided mannequin-based practice. Participants subsequently engaged in scenario-based exercises to perform the required skills. Facilitators assessed performance using standardized checklists.

### Study Instruments

Knowledge assessment questionnaires and skill checklists were developed in consultation with 2 independent emergency medicine experts and were validated on a small cohort of participants. The assessment included four sections: demographics (gender, age, education, occupation); confidence (prior first aid training experience); knowledge (13 multiple-choice questions covering core first aid topics such as chest compression rate and depth during CPR, CPR initiation criteria, bleeding control, proper splint application for fractures, communication with bystanders and emergency services during an incident, and evaluation for absence of breathing); and participants’ fears or concerns regarding first aid performance. Additionally, participants completed a self-efficacy checklist to rate their confidence and skill improvements before and after training. Participant feedback was collected post-training to guide improvements for future workshops.

### Assessment

Participants’ knowledge and self-efficacy were evaluated at two time points: pre-training (pre-test) and immediately post-training (post-test). A subset of participants completed a follow-up assessment at 3 months (re-test). During training sessions, facilitators presented simulated emergency scenarios such as drowning, poisoning, and road traffic accidents and asked participants to demonstrate key first aid skills on mannequins or volunteers for formative feedback. Psychomotor performance during these scenarios was observed but not systematically documented for analysis due to inconsistent form completion across stations and reliance on single rather than paired raters. Each participant was given up to three attempts per skill, each observed for at least two minutes. Immediate feedback was provided for any errors. The participant-to-facilitator ratio ranged from 5:1 to 10:1.

### Primary Outcomes

The primary outcomes were:

1. Change in participants’ first aid knowledge, determined by responses before and immediately after training
2. Change in participants’ self-efficacy across first aid domains, determined by validated instruments before and immediately after training

An exploratory secondary outcome was knowledge retention at 3 months post-training in a subset of participants.

### Statistical Analysis

Data were analyzed using R Studio version 2025.05.1+5137. Normality was assessed using the Shapiro-Wilk test. Internal consistency of the knowledge assessment was evaluated using Cronbach’s alpha, recognizing that the instrument was designed to assess heterogeneous first aid domains rather than a single unified construct. For self-efficacy scores, which demonstrated non-normal distribution post- intervention, Wilcoxon signed-rank tests were employed for paired comparisons. Knowledge assessment questions were analyzed using McNemar’s test for paired data. Overall knowledge scores were compared using paired t-tests, and Cohen’s d was calculated to determine effect size (small: d = 0.2, medium: d = 0.5, large: d = 0.8).

For the 3-month follow-up exploratory analysis, we compared post-training scores between the 15 participants who returned and the 53 who did not to assess potential selection bias. Retention rates were calculated as (3-month percentage / post-training percentage) × 100%. Bootstrap confidence intervals (10,000 replicates) were computed for retention estimates given the small sample size.

No correction for multiple comparisons was applied to the item-level McNemar’s tests (13 items) or the self-efficacy domain analyses (7 domains). Given the exploratory nature of this evaluation and the strong a priori rationale for expecting improvement across all assessed items following structured instruction, Bonferroni correction was considered overly conservative.

Descriptive statistics are presented as mean ± standard deviation (SD) for continuous variables and mean ± standard error of the mean (SEM) for figures, with frequencies and percentages for categorical variables. Statistical significance was set at p < 0.05. All tests were two-tailed.

The authors used Anthropic. (2026). Claude Opus 4.7 [Large language model]. https://claude.ai solely for grammatical editing and structural refinement of the manuscript text. The AI tool was not used for study design, data analysis, interpretation of results, or generation of scientific content. All outputs were reviewed and edited by the authors, who take full responsibility for the final manuscript.

### Ethical Considerations

Ethical approval for the study was obtained from the Aga Khan University Ethical Review Committee (ERC: 11605-36312). Written informed consent was secured from all participants prior to enrollment. Queries regarding the consent process were addressed by the principal investigator and the research team.

## Results

### Sociodemographics of the study participant*s*

Of the 132 trained participants, 68 had complete paired knowledge data and 100 had complete paired self-efficacy data. Among the knowledge cohort, the mean age was 25.0 ± 6.5 years with a range of 19–55 years. The majority of participants were female (n = 61, 89.7%), and most were single (n = 59, 86.8%), reflecting the younger composition of the sample.

In terms of education, 56.7% (n = 38) of participants held a bachelor’s degree or equivalent qualification (calculated among the 67 participants with non-missing education data), 19.4% (n = 13) held a master’s degree, 13.4% (n = 9) had completed education up to the intermediate level, and 10.4% (n = 7) had doctoral-level education.

Students formed the largest occupational group (60.3%, n = 41), followed by research assistants (25.0%, n = 17), and others, including post-doctoral fellows, technicians, and administrative staff (11.8%, n = 8). Approximately one-third of participants (36.8%, n = 25) reported having received prior first aid training.

Based on departmental affiliation data available for 72.1% (n = 49) of participants, the majority were from health-related programs (69.1%, n = 47). This represents a strategically important health-adjacent academic population: individuals likely to be first at scene in campus emergencies, informal responders in their households, and members of the future healthcare workforce. While the substantial proportion without prior first aid training (63.2%, n = 43) represented an opportunity to enhance emergency response competencies, the findings should be interpreted within the context of this specific population rather than extrapolated to the broader general community.

To further evaluate potential selection bias arising from incomplete assessment submission, baseline characteristics of participants who completed both pre- and post-training knowledge assessments (n = 68) were compared against those who submitted a form but did not complete both assessments (n = 26), yielding a total analytic sample of 94 participants (Supplementary Table S1). Three statistically significant differences were identified. Non-completers were significantly younger than completers (mean 22.3 ± 4.3 vs 25.0 ± 6.5 years; p = 0.027) and were more likely to report prior first aid training (61.5% vs 36.8%; p = 0.038). Education profiles also differed significantly (p = 0.029): non-completers had a higher proportion of intermediate-level education (38.5% vs 13.4%), while completers had a greater proportion of postgraduate qualifications (29.8% vs 11.5% Masters or PhD combined). Gender distributions were similar between groups (80.8% vs 89.7% female; p = 0.302). These findings suggest that non-completers represented a younger, more previously-trained subgroup with a lower educational level. Knowledge gains reported for the n = 68 completer cohort should therefore be interpreted with this compositional difference in mind.

### Self-Efficacy of participants

Participants demonstrated significant improvements in self-efficacy scores across all assessed domains following the intervention (Table 2). Mean scores for scene safety and emergency response readiness increased from 38.56 ± 2.55 before the workshop to 89.38 ± 1.29 after (p < 0.0001, Wilcoxon signed-rank test). Similarly, notable increases were observed in assessment skills (41.70 ± 2.78 to 83.19 ± 1.92), CPR skills (51.00 ± 3.10 to 93.96 ± 1.07), bleeding control (54.15 ± 3.15 to 92.01 ± 1.68), fracture and immobilization management (49.54 ± 2.86 to 89.44 ± 1.53), choking management (31.94 ± 2.71 to 86.98 ± 1.50), and motivation to prepare a first aid kit (35.05 ± 2.51 to 85.03 ± 1.70), all showing highly significant improvements (p < 0.0001 for all domains).

**Table 1.** Demographic characteristics of participants who completed the First Aid Training Workshop.

| Characteristic | n (%) or Mean ± SD |
| --- | --- |
| Age, years | 25.0 ± 6.5 |
| Range | 19–55 |
| **Gender** |  |
| Female | 61 (89.7) |
| Male | 7 (10.3) |
| **Marital Status** |  |
| Single | 59 (86.8) |
| Married | 9 (13.2) |
| **Education Level** |  |
| Up to Intermediate | 9 (13.4) |
| Bachelors or Equivalent | 38 (56.7) |
| Masters or Equivalent | 13 (19.4) |
| PhD or Equivalent | 7 (10.4) |
| Missing education | 1 (1.4) |
| **Occupation** |  |
| Student | 41 (60.3) |
| Research Assistant | 17 (25.0) |
| Post-doctoral Fellow | 3 (4.4) |
| Other (Technician/Admin/Clinical) | 5 (7.4) |
| Missing occupation | 2 (2.9) |
| **Prior First Aid Training** |  |
| Yes | 25 (36.8) |
| No | 43 (63.2) |

Values are presented as mean ± standard deviation for continuous variables and n (%) for categorical variables. N = 68 for knowledge assessment participants.

**Table 2.** Self-efficacy scores (0–100 scale) measured before and immediately after the training intervention across seven first aid skill domains.

| **Skill Category** | **Before Training**<br>**Mean ± SEM** | **After Training**<br>**Mean ± SEM** | **Change** | **P-value** |
| --- | --- | --- | --- | --- |
| **Scene Safety & Emergency Response** | 38.56 ± 2.55 | 89.38 ± 1.29 | +50.82 | <0.0001 |
| **Assessment Skills** | 41.70 ± 2.78 | 83.19 ± 1.92 | +41.49 | <0.0001 |
| **CPR Skills** | 51.00 ± 3.10 | 93.96 ± 1.07 | +42.96 | <0.0001 |
| **Bleeding Control** | 54.15 ± 3.15 | 92.01 ± 1.68 | +37.86 | <0.0001 |
| **Fracture & Immobilization** | 49.54 ± 2.86 | 89.44 ± 1.53 | +39.90 | <0.0001 |
| **Choking Management** | 31.94 ± 2.71 | 86.98 ± 1.50 | +55.04 | <0.0001 |
| **Motivation (First Aid Kit)** | 35.05 ± 2.51 | 85.03 ± 1.70 | +49.98 | <0.0001 |

*N = 100. Scores range from 0-100. Wilcoxon signed-rank test (two-tailed). SEM = Standard Error of the Mean.*
Values are presented as mean ± standard error of the mean (SEM). All domains showed highly significant improvement ( $p < 0.0001$ , Wilcoxon signed-rank test, two-tailed). $N = 100$ participants. The assessed domains included Scene Safety and Emergency Response Readiness (ensuring scene safety before approaching an unconscious or bleeding person; staying calm during an emergency involving resuscitation or bleeding control; recalling ambulance numbers in times of emergency; and timely activation of ambulance services), Assessment Skills (assessing level of consciousness; identifying when to stop cardiopulmonary resuscitation [CPR]; and recognizing life-threatening bleeding), CPR Skills (performing high-quality CPR; maintaining an adequate chest compression rate of 100–120 beats per minute; achieving appropriate compression depth of approximately 2 inches; correct hand placement during compressions; and allowing full chest recoil after each compression), Bleeding Control Skills (appropriately managing life-threatening bleeding; using protective barriers before providing care; applying direct pressure to the bleeding site; and performing wound packing for deep bleeding wounds), Fracture and Immobilization (correct positioning of splints; and appropriate splint application with safe transport considerations), and Choking Management (ability to manage choking in adults and infants).

The largest gains were observed in choking management (mean change: +55.04 points), scene safety and emergency response readiness (mean change: +50.82 points), and motivation to prepare a first aid kit (mean change: +49.98 points) and CPR skills (mean change: +42.96 points). Overall, the findings indicate a marked enhancement in participants’ self-efficacy regarding emergency response and first aid practices after the training intervention.

### Knowledge scores of participants at pre- and post- test

#### Internal Consistency of Knowledge Assessment

The first aid knowledge assessment was designed to assess breadth across heterogeneous first aid domains, including emergency response protocols, cardiopulmonary resuscitation techniques, bleeding control, fracture management, choking interventions, and clinical assessment skills rather than to measure a single underlying construct. Accordingly, internal consistency was low (Cronbach’s α = 0.443, 95% CI: 0.251–0.636), which is expected for a breadth-based assessment. Given this heterogeneity, we prioritized item-level analysis using McNemar’s test, with the total score serving as a secondary summary measure.

#### Overall Knowledge Change

A total of 68 participants completed both the pre- and post-training assessments of the First Aid Training Workshop. Overall knowledge scores improved significantly from 5.90 ± 2.01 points pre-training to 9.07 ± 2.42 points post-training (out of 13 possible points; paired t-test: p < 0.001), representing a mean gain of 3.18 ± 2.99 points. This improvement corresponded to a large effect size (Cohen’s d = 1.06); given the within-session pre–post design, this is best read as the upper bound of the immediate educational effect.

#### Item-Level Analysis

A comparison of pre- and post-test scores (Table 3; Figures 2–3) demonstrated substantial improvement in participants’ knowledge across most first aid domains following the educational intervention. Prior to training, correct response rates ranged from approximately 14.7% to 80.9%, indicating variable baseline knowledge of basic life support and first aid principles. After the training, post-test scores improved significantly, with most items showing substantial gains.

**Figure 2.**
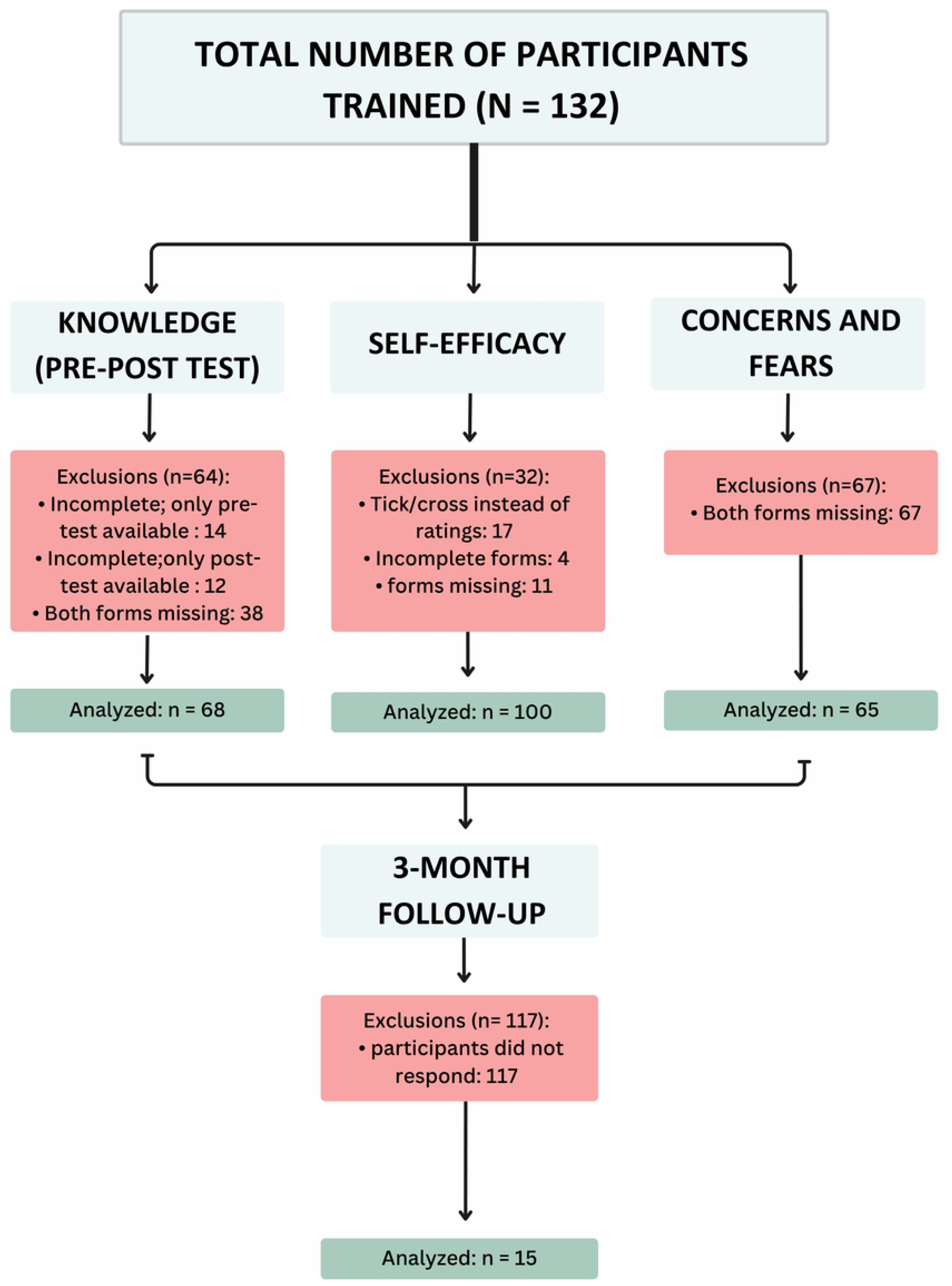
Bar chart comparing percentage of correct responses before (light gray bars) and after training (dark gray bars with diagonal hatching) for knowledge questions 1–7. P-values from McNemar’s test are displayed in white boxes above each question pair. Complete question stems are shown below each bar. All questions except Q1 and Q4 demonstrated significant improvement (p < 0.05). N = 68 participants.

**Figure 3.**
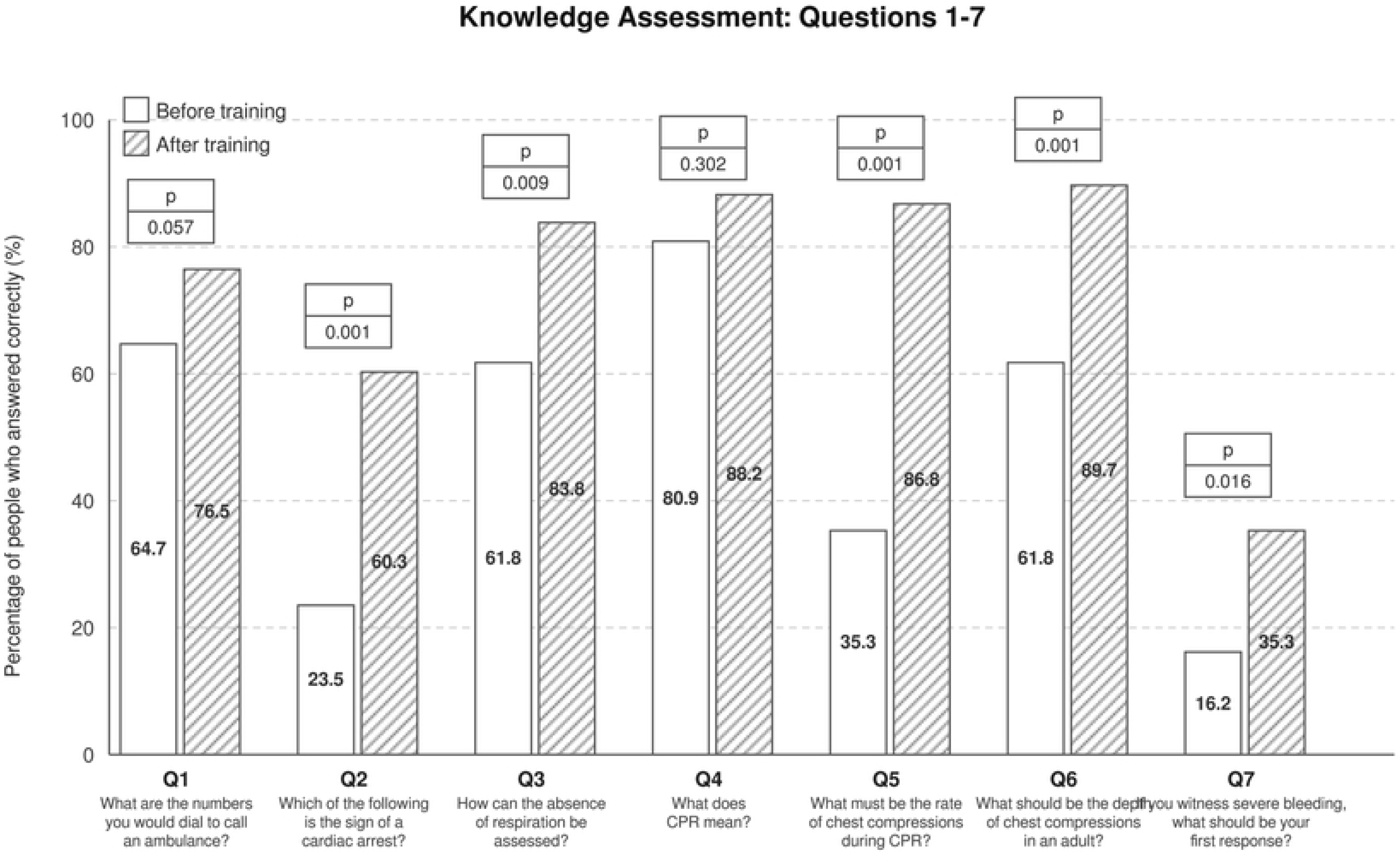
Bar chart comparing percentage of correct responses before (light gray bars) and after training (dark gray bars with diagonal hatching) for knowledge questions 8–13. P-values from McNemar’s test are displayed in white boxes above each question pair. Complete question stems are shown below each bar. Four of six questions demonstrated significant improvement (p < 0.05). Questions 11 and 12 showed non-significant improvements, likely due to high baseline knowledge (ceiling effect). N = 68 participants.

**Table 3.** Percentage of participants answering correctly for each of 13 knowledge questions, assessed before and immediately after training.

| Question | Pre-Training % | Post-Training % | Change (pp) | P-value<sup>a</sup> |
| --- | --- | --- | --- | --- |
| 1. What are the numbers to call an ambulance? | 64.7 | 76.5 | +11.8 | 0.057 ns |
| 2. Which is the sign of cardiac arrest? | 23.5 | 60.3 | +36.8 | <0.001*** |
| 3. How can absence of respiration be assessed? | 61.8 | 83.8 | +22.1 | 0.009** |
| 4. What does CPR mean? | 80.9 | 88.2 | +7.4 | 0.302 ns |
| 5. What is the rate of chest compressions during CPR? | 35.3 | 86.8 | +51.5 | <0.001*** |
| 6. What is the depth of chest compressions in adults? | 61.8 | 89.7 | +27.9 | <0.001*** |
| 7. First response to severe bleeding? | 16.2 | 35.3 | +19.1 | 0.016* |
| 8. How to give back blows to choking adult? | 35.3 | 77.9 | +42.6 | <0.001*** |
| 9. When placing splint on fracture, include? | 25.0 | 44.1 | +19.1 | 0.037* |
| 10. First response to cardiac arrest? | 14.7 | 64.7 | +50.0 | <0.001*** |
| 11. Why cover hands before approaching bleeding? | 75.0 | 85.3 | +10.3 | 0.121 ns |
| 12. Why is effective communication important? | 57.4 | 58.8 | +1.5 | 1.000 ns |
| 13. First question ambulance service asks? | 38.2 | 55.9 | +17.6 | 0.025* |

<sup>a</sup>N = 68. McNemar's test for paired binary data. pp = percentage points. \*p < 0.05, \*\*p < 0.01, \*\*\*p < 0.001.
Statistical significance determined by McNemar's test for paired binary data. Significance levels: \*\*\* p < 0.001, \*\* p < 0.01,
\* p < 0.05, ns = not significant. N = 68 participants.

The greatest improvements were observed in questions assessing the rate of chest compressions during CPR (Question 5), which increased by 51.5 percentage points (from 35.3% to 86.8%, p < 0.001), and the first response to a witnessed cardiac arrest (Question 10; +50.0 percentage points, from 14.7% to 64.7%, p < 0.001). Similarly, substantial gains were recorded in knowledge related to how to give back blows to a choking adult (Question 8; +42.6 percentage points), recognizing signs of cardiac arrest (Question 2; +36.8 percentage points), and identifying the appropriate depth of chest compressions (Question 6; +27.9 percentage points), all of which showed statistically significant improvements (p < 0.001).

Participants also showed significant enhancement in their understanding of emergency response communication and scene safety. Assessment of respiration (Question 3) rose from 61.8% to 83.8% (+22.1 percentage points, p = 0.009), first response to severe bleeding (Question 7) improved from 16.2% to 35.3% (+19.1 percentage points, p = 0.016), splint placement on fracture (Question 9) increased from 25.0% to 44.1% (+19.1 percentage points, p = 0.037), and understanding of the first question an ambulance service asks (Question 13) improved from 38.2% to 55.9% (+17.6 percentage points, p = 0.025). These findings suggest that the training not only improved technical first aid knowledge but also strengthened participants’ readiness to coordinate with emergency medical services.

Four questions showed non-significant improvements: the definition of CPR (Question 4; 80.9% to 88.2%, p = 0.302), the numbers to call for an ambulance (Question 1; 64.7% to 76.5%, p = 0.057), the reason to cover hands before approaching a bleeding site (Question 11; 75.0% to 85.3%, p = 0.121), and the importance of effective communication during emergencies (Question 12; 57.4% to 58.8%, p = 1.000). The high baseline scores for Questions 4, 11, and 12 (80.9%, 75.0%, 57.4% respectively) likely created ceiling effects limiting significant improvement.

Overall, McNemar’s test results indicated that 9 of 13 questions (69.2%) showed statistically significant improvements (p < 0.05), demonstrating the pre-post improvement of the training program in enhancing first aid knowledge. The consistent post-intervention increase across the majority of knowledge items reflects a broad and meaningful educational impact, suggesting that the structured workshop was associated with substantial short-term improvement in first aid knowledge in this health-adjacent academic population.

#### Concerns and Fears

Participants’ concerns and fears related to performing first aid were assessed on a scale from 1 (lowest concern) to 10 (highest concern) before and after the training intervention (Table 4). Baseline concerns were moderate across all assessed domains (range: 4.0–4.5 out of 10). Following the training, concern levels remained relatively stable, with no statistically significant changes observed (all p > 0.05, Wilcoxon signed-rank test).

**Table 4.** Participants’ concerns and fears before and after first aid training.

| Concern/Fear<sup>a</sup> | Before Mean ± SD | After Mean ± SD | Change | P-value |
| --- | --- | --- | --- | --- |
| Fear of infection during CPR or first aid | 4.1 ± 2.8 | 3.7 ± 2.9 | -0.4 | 0.133 |
| Fear of injuring the victim | 4.4 ± 2.7 | 4.6 ± 3.0 | +0.2 | 0.847 |
| Fear of injuring oneself | 4.0 ± 2.8 | 3.7 ± 2.9 | -0.3 | 0.527 |
| Fear of legal consequences | 4.5 ± 3.0 | 4.5 ± 3.0 | 0.0 | 0.819 |
| Fear of not recognizing signs of true injury or cardiac arrest | 4.4 ± 3.0 | 4.4 ± 3.3 | 0.0 | 0.944 |
| Fear of panic/reluctance to perform lifesaving skills | 4.1 ± 2.8 | 4.3 ± 3.1 | +0.2 | 0.617 |

<sup>a</sup> $N = 65$ . Scores range from 1 (lowest concern/fear) to 10 (highest concern/fear). \*Wilcoxon signed-rank test.
P-value calculated using Wilcoxon signed-rank test.

Two concerns showed slight, non-significant decreases: fear of infection during CPR or first aid (4.1 to 3.7, p = 0.133) and fear of injuring oneself (4.0 to 3.7, p = 0.527). Four concerns showed slight, non-significant increases or no change: fear of injuring the victim (4.4 to 4.6, p = 0.847), fear of legal consequences (4.5 to 4.5, p = 0.819), fear of not recognizing signs of true injury or cardiac arrest (4.4 to 4.4, p = 0.944), and fear of panic or reluctance to perform lifesaving skills (4.1 to 4.3, p = 0.617).

The persistence of concerns despite improved knowledge and self-efficacy may reflect multiple interpretations. One possibility is heightened awareness of the complexities and responsibilities associated with providing first aid; a realistic understanding appropriate for developing competent first responders. Alternatively, the modest increases in some fear domains may indicate that training inadvertently heightened awareness of potential risks. Both interpretations warrant consideration in designing future training interventions that explicitly address legal protections and build psychological readiness alongside technical competence.

### Three-Month Knowledge Retention: Exploratory Analysis

A subset of 15 participants (22.1% of the analysed cohort) completed a 3-month follow-up assessment. Among the 8 questions assessed at follow-up, participants demonstrated strong overall retention, maintaining 96.2% of their post-training knowledge level on average (3-month mean performance: 62.5% correct).

Retention varied substantially by question. Five questions retained or exceeded post-training performance: ambulance numbers (Question 1; 100% correct, retention 130.8%), assessment of respiration (Question 3; 86.7% correct, 103.4% retention), severe bleeding response (Question 7; 46.7% correct, 132.2% retention), choking management (Question 8; 93.3% correct, 119.7% retention), and splint placement (Question 9; 46.7% correct, 105.8% retention). Two questions showed moderate retention: sign of cardiac arrest (Question 2; 46.7% correct, 77.4% retention) and CPR compression rate (Question 5; 60.0% correct, 69.2% retention). One question showed notably low retention: the first response to cardiac arrest (Question 10) declined to 20.0% correct (30.9% retention rate), despite recording one of the largest immediate post-training gains (+50.0 percentage points) (Table 5).

**Table 5.** Percentage of participants answering correctly at baseline (pre-training), immediately post-training, and at 3-month follow-up for eight questions assessed at all three time points.

| Question | Pre-Training % | Post-Training % | 3-Month % | Retention Rate<sup>a</sup> (%) |
| --- | --- | --- | --- | --- |
| Ambulance numbers | 64.7 | 76.5 | 100.0 | 130.8 |
| Sign of cardiac arrest | 23.5 | 60.3 | 46.7 | 77.4 |
| Assess respiration | 61.8 | 83.8 | 86.7 | 103.4 |
| CPR compression rate | 35.3 | 86.8 | 60.0 | 69.2 |
| Severe bleeding response | 16.2 | 35.3 | 46.7 | 132.2 |
| Choking management | 35.3 | 77.9 | 93.3 | 119.7 |
| Splint placement | 25.0 | 44.1 | 46.7 | 105.8 |
| Cardiac arrest first response | 14.7 | 64.7 | 20.0 | 30.9 |
| **Average** | — | — | **62.5** | **96.2** |

<sup>a</sup> $N = 15$ (22.1% of the analysed cohort). Retention rate = (3-month % / post-training %) × 100%. Post-training proportions fixed at full-cohort values ( $n = 68$ ). Rates $>100\%$ indicate improvement beyond post-training levels. Blue shading: continued learning (retention $>100\%$ ). Red shading: notably low retention

To assess potential selection bias in the 3-month follow-up cohort, baseline characteristics and immediate post-training knowledge scores of the 15 participants who returned were compared against the 53 who did not (Supplementary Table S2). No statistically significant differences were observed on any examined variable: returners and non-returners were comparable in age (mean 26.3 ± 10.2 vs 24.6 ± 5.1 years; Welch p = 0.547), gender distribution (80.0% vs 92.5% female; p = 0.175), education profile (p = 0.255), prior first aid training (40.0% vs 35.8%; p = 0.770), pre-training knowledge scores (6.13 ± 2.07 vs 5.83 ± 2.01; p = 0.619), and post-training knowledge scores (9.60 ± 1.18 vs 8.92 ± 2.66; Mann-Whitney U p = 0.554). Returners showed marginally higher post-training performance with less variance, but this difference did not reach statistical significance and with n=15 the comparison was underpowered; non-significance does not establish representativeness.

#### Three-Month Knowledge Retention: Bootstrap Analysis

Three-month knowledge retention rates with bootstrap 95% confidence intervals are presented in Supplementary Table S3. Percentile intervals were derived from B = 10,000 resamples of the 15-participant follow-up cohort and are reported in preference to parametric estimates given the sample size. Retention was generally high but varied meaningfully across the eight assessed items. Two items demonstrated credible retention of post-training gains with narrow confidence intervals: choking management (119.7%; 95% CI 102.6–128.3%) and assessment of respiration (103.4%; 95% CI 79.5–119.3%). Ambulance contact numbers showed perfect 3-month performance (100% correct, n=15), precluding confidence interval estimation but indicating complete retention of this ceiling-effect item. A further three items showed retention at or modestly above post-training levels with wider confidence intervals: severe bleeding response (132.2%; 95% CI 56.7–207.8%), splint placement (105.8%; 95% CI 45.3–166.2%), and sign of cardiac arrest (77.4%; 95% CI 33.2–121.6%). CPR compression rate showed below-post-training retention (69.2%; 95% CI 38.4–99.9%). The most notable departure was the first-response-to-cardiac-arrest item, which retained only 30.9% of its post-training level (95% CI 0.0–61.8%) despite one of the largest immediate gains recorded (+50.0 percentage points). The unweighted mean retention across items was 96.2% (95% CI 78.6–112.7%), best interpreted alongside item-level estimates given the heterogeneity in retention patterns. Consistent with the exploratory nature of these findings, returners and non-returners did not differ significantly on measured characteristics (Supplementary Table S2), though the modest power limitations mean upward bias in retention estimates cannot be formally excluded.

## Discussion

This single-centre, pre–post quasi-experimental evaluation examined changes in knowledge, self-efficacy, persistent concerns regarding first aid performance, and exploratory three-month knowledge retention following a structured multi-station workshop delivered to health-adjacent trainees in Karachi. The intervention was associated with substantial gains in both knowledge and self-efficacy across all assessed domains, while subjective concerns and fears remained a comparatively resistant target for short-format training. These findings inform the design of structured first aid education in this population while highlighting where current curricular models require enhancement.

### Demographic and Population Characteristics

The cohort was predominantly female (89.7%), young (mean age 25.0 ± 6.5 years), and educationally advantaged, with 56.7% holding a bachelor’s degree. This demographic profile mirrors that of comparable first aid training studies conducted regionally and globally, in which female participation and student or early-career representation typically dominate enrolment [15–17]. The over-representation of women is consistent with broader patterns of female engagement in community health interventions and the gendered structure of caregiving in South Asian households, in which women are disproportionately positioned as informal first responders during domestic emergencies [17]. The strategic value of training a health-adjacent academic population should not be underestimated: such individuals are simultaneously the future healthcare workforce, the most likely first-at-scene responders within campus environments, and the informal medical reference points for their families and neighbourhoods. Comparable rationales have underpinned community-based programs in Chicago, Cape Town, Iganga (Uganda), and Ibadan (Nigeria), where trained laypersons drawn from socially central groups have demonstrably extended the reach of early prehospital care [3,18–21]. Notably, 63.2% of our cohort reported no prior first aid training, underscoring an unmet need even within a relatively well-educated population in a megacity with comparatively better-developed services than rural Pakistan.

### Self-Efficacy Outcomes

Self-efficacy gains were the most striking single feature of the intervention. All seven assessed domains demonstrated highly significant improvement (p < 0.0001), with mean changes ranging from +37.86 points in bleeding control to +55.04 points in choking management and +50.82 points in scene safety and emergency response readiness. The magnitude and breadth of these gains are consistent with; and in several domains exceed those reported in international community-training evaluations. The Trauma Responders Unify to Empower (TRUE) community course in Chicago demonstrated significant self-efficacy gains across seven empowerment-based items, with effects sustained at six months [18]. Similarly, a structured CPR training initiative for Polish working adults produced significant pre–post improvements in self-confidence and willingness to perform resuscitation [9]. A systematic review by Riggs and colleagues encompassing more than 35,000 layperson participants confirmed that self-efficacy is weakly but consistently associated with CPR psychomotor skill level and that training-induced self-efficacy gains mediate willingness to intervene [22]. Bandura’s foundational construct of self-efficacy emphasizes that perceived capability is a critical determinant of whether knowledge translates into action; in the context of bystander emergencies, this construct is operationally inseparable from outcomes, as hesitation of even a few minutes can decrease the probability of survival from cardiac arrest by 7–10% per minute [23,24].

The largest gains in our cohort occurred in choking management, scene safety, and motivation to assemble a first aid kit; domains in which baseline self-efficacy was particularly low (31.94, 38.56, and 35.05 out of 100, respectively). This pattern suggests that the training addressed not merely technical instruction but also the cognitive scaffolding required to translate knowledge into intent: knowing which numbers to call, when to act, and how to prepare. Given that early activation of emergency medical services is the first and most consequential link in the chain of survival and is independently associated with up to a three-fold increase in survival, these findings carry direct public health relevance for Pakistan, where dispatch-assisted CPR remains rare and bystander activation is the dominant pathway to definitive care [9–11].

### Knowledge Outcomes

Overall knowledge scores improved from a mean of 5.90 ± 2.01 to 9.07 ± 2.42 out of 13 (paired t-test, p < 0.001; Cohen’s d = 1.06), corresponding to a large effect size that approaches or exceeds the magnitudes reported in comparable LMIC and university-based interventions. A recent quasi-experimental evaluation in Kazakhstan reported a significant pre–post knowledge gain among non-medical individuals receiving structured first aid training [25]. A landmark Karachi-based study of bystander CPR by Khan and colleagues observed an increase in mean knowledge scores from 47.8/100 to 70.2/100 immediately after training, with modest decay at six months, a pattern broadly comparable to ours [11]. The low Cronbach’s α (0.443) reported for our 13-item assessment was anticipated and reflects the breadth-based design of the instrument, which deliberately samples multiple, conceptually distinct first aid domains rather than measuring a single latent construct. Item-level analysis using McNemar’s test is therefore the appropriate primary inferential approach, with the summary score serving a complementary purpose [14].

Item-level findings are interpretable and clinically meaningful. The greatest improvements were observed for the rate of chest compressions during CPR (+51.5 percentage points), the appropriate first response to witnessed cardiac arrest (+50.0 percentage points), and back-blows for the choking adult (+42.6 percentage points). These domains correspond directly to the time-critical actions emphasized by international resuscitation guidelines and constitute the operational essence of the chain of survival [5,10]. Substantial improvements were also observed in emergency medical services communication skills, including knowledge of ambulance contact numbers and the structure of dispatcher questioning, which directly address the documented coordination failures responsible for delayed bystander response in Pakistan [11]. Conversely, four questions exhibited non-significant change:Ambulance numbers (Question 1), the definition of CPR (Question 4), rationale for hand protection prior to bleeding intervention (Question 11), and the importance of effective communication (Question 12). All four demonstrated high baseline scores (57.4–80.9%), generating a clear ceiling effect that constrained the statistical detectability of further gain rather than indicating a failure of pedagogy. Improvements in bleeding (Question 7, +19.1 percentage points) and splinting (Question 9, +19.1 percentage points), although statistically significant, were comparatively modest. This finding is consistent with the literature: first aid curricula frequently allocate proportionally more time to CPR than to trauma management, reflecting both the centrality of cardiac arrest in resuscitation pedagogy and the relative complexity of operationalizing trauma stabilization within a single workshop [8,14,26]. Recent systematic reviews of hemorrhage control training in LMICs explicitly call for greater curricular weight to be given to bleeding control and trauma stabilization, particularly in regions where penetrating injury and road traffic trauma dominate the prehospital epidemiology [8,26]. Our findings reinforce the case for rebalancing future iterations of similar workshops to provide comparable depth across cardiac and trauma domains.

### Concerns and Fears Regarding First Aid Provision

In contrast to the marked improvements in knowledge and self-efficacy, participants’ concerns and fears regarding the provision of first aid remained essentially unchanged after training. Baseline concerns were moderate across all six assessed domains (4.0–4.5 out of 10) and showed no statistically significant change following the intervention. Small, non-significant reductions in fear of infection and fear of injuring oneself suggest that scene safety and personal protection messaging was at least partially internalized, while small, non-significant increases in fear of injuring the victim and fear of panic suggest that training may have heightened participants’ realistic appraisal of the responsibility entailed in intervention. Notably, fear of legal consequences remained completely unchanged.

This pattern is of substantial interpretive and practical importance. Qualitative evidence from bystanders to road traffic accidents identifies legal exposure, fear of causing harm, and emotional unpreparedness as among the most consistent barriers to layperson intervention, even when technical competence has been demonstrably acquired [12, 27]. A recent psychometric study further confirms that fears related to blood, injection, and mutilation significantly reduce the self-reported likelihood of providing first aid, independent of training history [28]. The persistence of legal fear in our cohort is particularly pertinent to the Pakistani context, where; unlike the United States, Canada, the European Union, China, India, and the United Arab Emirates no comprehensive Good Samaritan statute currently protects bystanders from civil or criminal liability arising from good-faith emergency assistance [29–33]. Empirical evidence from comparable South Asian settings reinforces this concern: a recent peer-reviewed analysis of two nationwide Indian surveys identified police questioning, hospital detention, and upfront payment demands as the dominant deterrents to bystander assistance, with statutory codification of Good Samaritan protections subsequently associated with a 65.4% rise in willingness to assist [33]. Publicly documented incidents from Karachi indicate that identical patterns of hesitation persist in Pakistan in the absence of comparable legal cover [34]. Our findings therefore suggest that technical training alone cannot neutralise structurally generated fears; future first aid curricula in Pakistan should pair skill instruction with explicit discussion of the legal landscape and structured psychological preparation, drawing on intention-focused pedagogy of the kind advocated by Panchal and colleagues to close the gap between competence and willingness to act [35].

### Three-Month Knowledge Retention: Bootstrap Analysis

Exploratory analysis of three-month retention among the 15 returning participants indicated a mean retention of 96.2% of post-training knowledge levels (bootstrap 95% CI 78.6–112.7%). Retention was generally high but varied across items. Two items retained essentially all of their post-training gains: choking management (119.7%; 95% CI 102.6–128.3%) and assessment of respiration (103.4%; 95% CI 79.5–119.3%). Ambulance contact numbers showed perfect retention at three months (all 15 participants answered correctly; confidence interval not estimable), though this ceiling effect limits interpretation. Values exceeding 100% reflect sampling resolution (±6.7pp per respondent) and indicate complete retention; informal rehearsal cannot be excluded. Conversely, knowledge of the first response to witnessed cardiac arrest declined sharply, retaining only 30.9% of its post-training level (95% CI 0.0–61.8%) despite one of the largest immediate post-training gains recorded (+50.0 percentage points). This profile of rapid acquisition followed by rapid decay of action-sequencing knowledge is well documented in the CPR-retention literature. The systematic review by Riggs and colleagues concluded that CPR psychomotor skills typically deteriorate within three months and then plateau, with knowledge of action sequences particularly vulnerable to decay [22]. The earlier Karachi-based PLSP feasibility study by Khan et al. similarly reported a modest reduction in mean CPR knowledge from 70.2/100 to 66.5/100 at six months, alongside a significant decline in psychomotor skill components [11]. The Public Access and Tourniquet Training (PATTS) study by Goralnick et al. reported that although 87.7% of laypersons could correctly manage bleeding immediately after a one-hour course, only 54.5% retained this ability at three to nine months [26]. Taken together, these convergent findings indicate that first aid education is better conceived not as a one-time intervention but as a continuing programme structured around periodic refresher cycles, ideally every three to six months for skill components and six to twelve months for cognitive components [22,36,37].

These retention estimates should be interpreted in the context of a modest three-month follow-up rate of 22.1% (n = 15 of 68). As documented in Supplementary Table S2, a sensitivity analysis found no statistically significant differences between returners and non-returners on any measured characteristic, including age, gender, education, prior training, and pre- and post-training knowledge (Mann-Whitney U p = 0.554 for post-training scores). This reduces, but does not eliminate, concern about selection bias: returners scored modestly higher post-training (9.60 ± 1.18 vs 8.92 ± 2.66), and although this difference was not statistically distinguishable at the available sample size, the limited power of the comparison means a slight upward bias cannot be formally excluded; particularly as follow-up cohorts in training evaluations tend to over-represent more engaged, higher-performing participants [38]. The retention estimates derived from this cohort may therefore represent modest upper-bound values. We addressed this explicitly through transparent sensitivity analysis and bootstrap interval estimation, which we regard as methodological strengths of the evaluation. With these caveats, the broadly high retention observed across multiple knowledge domains is consistent with durable cognitive learning from structured training in this population, while the sharp decay of the cardiac-arrest response item underscores the need for periodic reinforcement of time-critical procedural skills.

### Public Health and Educational Implications

The findings of this study carry implications for first aid education in Pakistan and comparable LMIC settings, with each implication scaled to what a single-centre, pre–post evaluation can reasonably support. First, the magnitude of knowledge and self-efficacy gains observed here is consistent with the international evidence base for structured multi-station first aid instruction and supports the integration of such workshops into university curricula and professional development programmes for health-adjacent trainees. Analogous integration is well established in several high-income settings, where school-based basic life support and first aid education are embedded within public health and educational frameworks, including longstanding curriculum-based programmes in Norway and regions of the United States [39]. Pakistan’s PLSP initiative offers a ready vehicle for further institutional uptake, and our results provide locally generated empirical support for its core curricular content within trainee populations [11,13]. Whether comparable gains are achievable in unselected community samples, the broader target of community bystander training, will require evaluations with explicitly community-recruited cohorts.

Second, the population reached here, health-adjacent trainees drawn from health-related programs across Karachi, occupies a strategically useful position within the chain of survival. As future healthcare professionals, informal household responders, and potential trainers of subsequent cohorts, they represent a high-yield group for early investment, though one that should be understood as complementing rather than substituting for general-population reach.

Third, the persistence of fears in our cohort, particularly fear of legal consequences, which did not shift despite substantial gains in knowledge and self-efficacy, underscores the parallel need for legislative reform. The introduction of a Pakistan Good Samaritan statute, currently under public discussion, would directly address one of the most consistently documented barriers to bystander intervention and would complement educational interventions of the kind evaluated here [29–34].

### Strengths

This study has several notable strengths. First, to our knowledge, it is among the first evaluations in Pakistan to simultaneously assess knowledge, self-efficacy, fears and concerns, and three-month retention within a single intervention cohort. This multi-domain framework provides a more complete picture of training impact than the predominantly CPR-focused, knowledge-only evaluations that have dominated the regional literature.

Second, the study used established statistical methods suited to each outcome. These were McNemar’s test for paired binary items, Wilcoxon signed-rank tests for non-normal continuous outcomes, paired t-tests with Cohen’s d effect-size estimation for normally distributed summary scores, and bootstrap percentile confidence intervals for small-sample retention estimates. This choice of methods is appropriate to the field.

Third, recognising the heterogeneous nature of the knowledge instrument, the manuscript transparently reports Cronbach’s α and justifies the prioritization of item-level analysis, an approach increasingly recommended in first aid education research.

Fourth, sensitivity analyses comparing completers to non-completers and returners to non-returners were undertaken and transparently reported, giving the reader the information needed to contextualize potential selection bias.

Finally, the intervention was delivered through six skill-based stations led by certified facilitators, with mannequin practice and scenario-based exercises. This design follows pedagogical best practice and is reproducible within the resource constraints typical of LMIC academic medical settings.

### Limitations and Practical Constraints

This evaluation has several limitations. First, the analytic sample for knowledge assessment (n = 68) was smaller than the trained cohort (n = 132) owing to partial loss of paper-based forms and incomplete paired assessments. The surviving sample remained adequate to detect the large observed effect (Cohen’s d = 1.06), and the parallel self-efficacy sample (n = 100) preserved power for the principal affective outcome. Future iterations will migrate to digital data capture with dedicated form-completion oversight.

Second, the three-month follow-up rate of 22.1% (n = 15 of 68 in the knowledge cohort) is consistent with attrition reported in comparable community-based studies but limits the generalisability of retention estimates. We addressed this through bootstrap confidence intervals and explicit comparison of returners and non-returners. As no significant differences emerged between the two groups, the resulting estimates are presented as exploratory values that may represent a modest upper bound. Future cycles will incorporate scheduled reminders, modest participation incentives, and pre-arranged assessment appointments.

Third, psychomotor skills were observed and remediated in real time during scenario-based exercises but were not included in the formal analytic dataset because of inconsistent station-level documentation and reliance on single rather than paired raters. Standardised digital checklists and inter-rater reliability protocols are planned for the next iteration.

Fourth, the knowledge questionnaire was developed with two emergency medicine experts and pilot-tested locally rather than benchmarked against an externally validated instrument, reflecting the current absence of validated Urdu-language or South Asian first aid assessment tools. Future studies will pair locally adapted items with validated international instruments to enable cross-study comparison.

Fifth, knowledge and self-efficacy were measured immediately before and immediately after the intervention within the same session. This design is susceptible to demand characteristics, in which participants infer the trainers’ expectations and respond accordingly, and to testing effects, in which pre-test exposure primes recognition of correct responses on the post-test. Self-reported self-efficacy is particularly vulnerable to both. The immediate post-workshop estimates should therefore be read as upper bounds on training-related gain rather than as unbiased effect sizes. Future iterations will incorporate a delayed post-test administered outside the training session and, where feasible, a non-trained comparison cohort to help separate training-related change from session-related artefact.

Finally, the cohort is predominantly female, young, and drawn from a health-adjacent academic environment. The findings should be read as applicable to this specific subgroup, which is itself a strategically important segment of the future health workforce and informal first-responder network, rather than extrapolated to the wider Pakistani population.

## Conclusion

In this single-centre, single-arm, pre–post evaluation, a structured multi-station first aid workshop produced large gains in knowledge and self-efficacy among health-adjacent trainees in Karachi, while concerns about legal exposure, harming the victim, and panic remained essentially unchanged. The persistence of these fears despite clear improvement in technical knowledge is perhaps the most consequential finding of the study: it indicates that the bystander-action gap in Pakistan is as much structural and psychological as it is educational, and that skills instruction in isolation is unlikely to close it.

Two implications follow. At the curricular level, future iterations of similar workshops should integrate explicit content on the legal landscape facing bystanders in Pakistan, scenario-based psychological preparation for the emotional reality of intervention, and intention-focused pedagogy designed to convert confidence into willingness to act. At the policy level, the persistence of legal fear even among health-adjacent participants who might be expected to have some baseline familiarity with the medico-legal environment reinforces the case for a Pakistan Good Samaritan statute comparable to those already enacted in the United States, the European Union, the United Arab Emirates, and, most directly relevant, India, where statutory codification was followed by a substantial rise in willingness to assist.

By jointly examining knowledge, self-efficacy, fears, and exploratory medium-term retention within a single intervention cohort, this evaluation broadens the evidence base for first aid training in Pakistan beyond the CPR-knowledge focus that has dominated the regional literature, and supports the continued use of the multi-station workshop format as a building block for larger national initiatives such as the Pakistan Life Savers Programme. The next phase of this work will extend recruitment beyond health-trainee populations, incorporate standardised psychomotor assessment, and use a delayed-training or stepped-wedge design to permit causal inference.

## Data Availability

The de-identified dataset underlying this study is provided as Supporting Information (S1 Dataset)

## Acknowledgements

The authors would like to acknowledge the Pakistan Life Savers Programme (PLSP) for its invaluable support in the development and delivery of the workshop. We are especially grateful to Dr. Noor Baig for his guidance and mentorship throughout the project. We also extend our sincere thanks to the trainers who contributed to participant instruction and skills development.

## Data Sharing Statement

The de-identified dataset underlying this study is provided as Supporting Information (S1 Dataset)

## Author contributions

**MRAJ** contributed to data curation, methodology development, investigation activities, resource acquisition, and writing of the original draft of the manuscript. **AA** contributed to conceptualization of the study, project administration, resource provision, investigation activities, and manuscript review and editing. **IN** contributed to investigation activities and writing of the original draft of the manuscript. **SHS** contributed to data curation, formal analysis, preparation of the results section, and manuscript writing. **YS** contributed to supervision of the project, provision of academic guidance, and manuscript review and editing. **MF, AS,** and **SI** contributed to data extraction, project administration, and investigation activities. **AP** served as a senior supervisor and contributed to project guidance, supervision, manuscript review and editing, and final approval of the manuscript.

## Supporting information

**Supplementary Table S1.** Baseline characteristics of participants who completed both pre- and post-training knowledge assessments (n=68) versus those who submitted a form but did not complete both assessments (n=26).

**Supplementary Table S2.** Baseline and post-training characteristics of participants who completed the 3-month follow-up assessment (n=15) versus those who did not return (n=53), within the knowledge cohort (N=68).

**Supplementary Table S3.** Three-month knowledge retention rates with bootstrap 95% confidence intervals.

